# A low molecular weight dextran sulphate, ILB®, for the treatment of amyotrophic lateral sclerosis (ALS): an open-label, single-arm, single-centre, phase II trial

**DOI:** 10.1101/2023.08.27.23294691

**Authors:** Venkataramanan Srinivasan, Victoria Homer, Darren Barton, Abigail Clutterbuck-James, Siân Jenkins, Claire Potter, Kristian Brock, Ann Logan, Donna Smith, Lars Bruce, Zsuzsanna Nagy, Simon P Bach

**Author notes:** Corresponding author contact details: Dr Venkataramanan Srinivasan, Neurology, Queen Elizabeth Hospital, University Hospitals Birmingham NHS Foundation Trust (UHBFT), Birmingham. B15 2WB. United Kingdom.

## Abstract

**Background:** Amyotrophic lateral sclerosis (ALS), also known as Lou Gehriǵs disease, is a rare neurological disease and is the most common motor neurone disease. It is a fatal disease with specific loss of motor neurons in the spinal cord, brain stem, and motor cortex leading to progressive paralysis and usually death within five years of diagnosis. There remains no cure for ALS, and management is focused on a combination of neuroprotective medication, respiratory support, and management by multidisciplinary clinics.

**Patients and Methods:** This prospective, single-arm, open-label phase II clinical trial of sustained weekly administration of 2 mg/kg ILB® (a low-molecular weight dextran sulphate) was conducted in a single UK hospital. Eligible patients were at least 18 years and had a definite diagnosis of ALS according to El Escorial Criteria. The co-primary outcomes were safety, tolerability, and quantity of ILB® administered. All evaluable patients were analysed in a modified intention-to-treat analysis.

EudraCT number. 2018-000668-28

**Findings:** Between 18-Apr-2019 and 27-Mar-2020, 11 patients were recruited and treated for up to 38 weeks. There were no treatment terminations or withdrawals. One serious adverse event was reported, which was not related to ILB® and resolved without sequalae. 270 mild/moderate adverse events were reported with no intolerable events occurring during the trial. The total number of ILB® treatments administered per patient ranged from 4 to 38, with a cumulative dose ranging from 745 to 6668 mg. As a result of the COVID-19 pandemic and the high-risk status of study participants, recruitment and treatment was suspended early in Mar-2020. At the long-term follow-up, three patients had died after the trial was halted, between 53 and 62 weeks after their final ILB® injection.

**Interpretation:** Long-term weekly ILB® injections of 2 mg/kg was well tolerated and had an acceptable safety profile in patients with ALS.

## Introduction

Amyotrophic lateral sclerosis (ALS), also known as Lou Gehriǵs disease, is a rare neurological disease and the most common motor neurone disease (MND). It is fatal, with specific loss of motor neurons in the spinal cord, brain stem and motor cortex leading to progressive paralysis and usually death within five years of diagnosis. There is no cure for ALS. Management is focused on a combination of neuroprotective medication, respiratory support, and management by multidisciplinary clinics [1]. Patients treated by ALS care teams may have higher quality of life (QoL) and longer survival as malnutrition and dehydration are common as ALS advances [2, 3]. Later clinical management often includes percutaneous gastric feeding and non-invasive ventilation-based support.

Only one commonly used medication, Riluzole licensed in 1996, has been proven to prolong survival in patients with ALS [4]. Riluzole possesses anti-glutamatergic properties that reduce excitotoxicity in ALS. Randomised controlled trials have demonstrated that Riluzole slows disease progression [5–7] and prolongs survival in patients with ALS by about 2-3 months [4]. More recently, Edaravone and the combination of sodium phenylbutyrate (PB) and taurursodiol called AMX0035 or Relyvrio have received FDA approval as alternative ALS therapeutics that may also modestly prolong life [8]. However, numerous trials have been unable to identify a curative or disease-modifying agent. Therefore, research has focussed on slowing disease progression by targeting known pathophysiological pathways or genetic defects.

The mechanisms of neuronal death in ALS are yet to be fully understood, but multiple mechanisms appear to contribute to the disease development and progression [9, 10]. Treatments targeted to a single pathogenic mechanism are not a feasible option for the very heterogeneous sporadic ALS population, however, promotion of growth factor dependent survival of affected neuronal populations offers a promising therapeutic avenue [10, 11]. As a potent survival-promoting factor for motor neurons, hepatocyte growth factor (HGF) has been suggested as an ALS treatment [12, 13]. HGF is shown to reduce motor neuron death and axonal degeneration, prolonging life in transgenic ALS mouse models [14], attenuate spinal motor neuron degeneration in transgenic ALS rat models [15], and improve functional recovery in a non-human primate model of contusive cervical spinal cord injury [16].

Dextran sulphate, originally developed and explored for its anticoagulant properties, has additional clinical effects, with low molecular weight dextran sulphate (LMW-DS) formulations investigated clinically as antiviral compounds in patients with HIV-1 [17, 18], and as medicines to treat those with post-perfusion lung damage [19], and stroke [20]. ILB® is a LMW-DS. This molecule was found to be neuroprotective in vitro and in animal models [21]. Detailed mechanism of action studies indicate that this neuroprotective effect is due to the ability of ILB® to mobilise heparin binding growth factors, such as HGF, and cytokines from the endothelial bed [21, 22]. Administration of ILB® at a wide range of concentrations has been shown to be well-tolerated and leads to a dose-dependent increase in plasma HGF in human healthy volunteers [22–24]. Furthermore, a phase II open-label pilot clinical study demonstrated safety and tolerability of short-term ILB® administration (five weekly doses of 1 mg/kg) in patients with ALS with improvements in functional and biochemical parameters while receiving treatment [25].

Recent reports have demonstrated a complex pleiotropic mechanism of action of ILB® and indicated its potential as a novel neuroregenerative medicine [21]. Given these data, the aims of this trial were to assess the safety and tolerability of longer-term and higher dose ILB® treatment in patients with ALS.

## Patient and Methods

### Study design

The ALS trial was a single-arm, open-label, phase II clinical trial recruiting participants from a motor neuron disease clinic at the Queen Elizabeth Hospital, Birmingham UK. As a first-in-patient study evaluating longer-term and higher ILB® dosing in patients with ALS, the first two recruits were considered as sentinel patients. These patients were recruited in series and safety data ascertained prior to expanding recruitment.

Ethical approval for the trial protocol (ultimately Version 8.0 dated 25-Nov-2020) was obtained from the South Central - Oxford B Research Ethics Committee and the local institutional review board and ethical committee in accordance with national and international guidelines. The current version of the protocol and patient information sheets are included in appendices S1 and S2.

### Patients

Eligible patients were ≥18 years and had a definite diagnosis of ALS according to El Escorial Criteria [26]. All patients demonstrated either a presence of Upper Motor Neuron (UMN) as well as Lower Motor Neuron (LMN) signs in the bulbar region and at least two of the other spinal regions (cervical, thoracic or lumbosacral), or a presence of UMN and LMN signs in all three spinal regions (cervical, thoracic or lumbosacral).

Patients had a forced vital capacity (FVC) ≥50% of predicted value for gender, height, and age at screening and/or a mean Sniff Nasal Inspiratory Pressure (SNIP) ≥50% of predicted value for age, with adequate haematological function, an International Normalised Ratio (INR) ≤1.5, Activated Partial Thromboplastin Time (aPTT) ≤40 seconds, and a Prothrombin Time (PT) ≤13.5 seconds.

Patients were excluded who required radiologically inserted gastrostomy or percutaneous endoscopic gastroscopy feeding, or who had active peptic ulcer disease, or abnormal liver function (defined as aspartate transaminase (AST) and/or alanine transaminase (ALT) >3 times upper limit of normal) or uncontrolled severe hypertension, or any head trauma, or who had undergone intracranial or spinal surgery within three months of trial entry, or had a diagnosis of a pulmonary illness or another neurodegenerative disease, or who were using an anticoagulant/low molecular weight subcutaneous heparin, or had evidence of major psychiatric illness.

Pregnant and breast-feeding women were excluded and those with reproductive potential were required to use effective methods of contraception. All patients gave written informed consent.

Patient registration into the trial by the treating clinician was by telephone to the central registration service at the Cancer Research UK Clinical Trials Unit (CRCTU) at the University of Birmingham.

### Procedures

Following successful screening, and prior to initiation of ILB® therapy, Riluzole, if taken, was stopped. A minimum 28-day washout period prior to ILB® administration was mandated to avoid potential drug interactions, given that Riluzole may inhibit elements of the mechanism of action of ILB® [21, 25].

ILB® was administered at a dose of 2 mg/kg (in a 100 mg/mL formulation) *via* subcutaneous injections, given as a single injection in alternating sides of the abdomen, once per week for 10 weeks on an out-patient basis. Dosing beyond 10 weeks, initially to 24 weeks, and subsequently to a maximum of 48 weeks was accompanied by a formal review at the time of extension, and subsequently every 90 days to determine whether they continued to meet eligibility criteria, and the most suitable treatment options. During these discussions a joint decision between the patient and their treating clinician was made as to whether the patient continued to receive trial treatment.

Adverse events (AEs) according to NCI-CTCAE v4.0 [27] were recorded from trial registration until 30 days after final treatment. All liver related toxicities with ALT >3x upper limit of normal (ULN) and total bilirubin >2x ULN (>35% direct), or ALT >3x ULN and INR >1.5 were reported as serious adverse events (SAEs). In addition, ILB® administration was terminated if a patient experienced an SAE assessed to be related to ILB® or an SAE that required discontinuation of ILB®; a decrease in Revised ALS Functional Rating Scale (ALSFRS-R) score >50% compared to baseline [28]; abnormal coagulation defined as PTT or aPTT >1.5 x ULN, or INR >1.8x ULN. ILB® administration was also stopped if a patient required anticoagulant or low molecular weight subcutaneous heparin medication, or if a patient exhibited a decrease in ALSFRS-R score of >50% compared to baseline during the clinical trial period.

Study participants were closely monitored for drug induced liver injuries (DILIs). Schedules for phase II liver chemistry monitoring and required follow up assessments are presented in supplementary materials (Table S3).

The ALS Functional Rating Score (ALSFRS-R) [28] was assessed at the initial screening visit and at each subsequent visit prior to treatment or during each follow-up, combined with the ALS Assessment Questionnaire (ALSAQ-40) [29] to monitor symptom progression and quality of life (QoL), respectively. The ALSFRS-R was completed by the treating clinician, with the ALSAQ-40 completed independently by patients. Following premature closure of the trial due to the COVID-19 pandemic, all patients were invited to provide QoL data during a single remote follow-up visit (all within eight weeks of each other; 15 months following the closure of the trial). Patients were re-consented electronically, and the visit was conducted *via* video conference with the ALS study clinician to assess ALSFRS-R, ALSAQ-40, and concomitant medication, including any current ALS treatment. The onset-related data was collected by the research team from the participants.

Blood and urine samples were collected for biochemical, pharmacokinetic and biomarker assessments; see Sample Collection section.

### Outcomes

The co-primary outcomes of this trial used to assess tolerability and safety of ILB® were: the quantity of study drug administered defined as total drug administered and number of administrations; the number and length of treatment interruptions plus number of discontinuations; the incidence of SAEs and AEs using CTCAE v4.0 [27], and the incidence of ‘intolerable’ AEs, defined as satisfying all of the following:

- Grade 3, 4 or 5 AE according to CTCAE v4.0
- Rated by the investigator as being possibly, probably, or definitely related to ILB®
- Either rated as a SAE or, if an AE, resulting in discontinuation of ILB® > 3 weeks.

Secondary outcome measures included description of the effect of ILB® on the severity of ALS symptoms already present at trial entry, the development of new ALS-associated symptoms, and the QoL of patients as determined by ALSFRS-R and ALSAQ-40. These questionnaires are in the form of ordinal scales that assess communication, mobility, feeding, dressing and respiration; and the subjective well-being of patients including emotional well-being [28, 29]. Collected questionnaire data was transformed to derive summary scores and scales. Derivation was performed in accordance with the respective user manuals.

Pre-specified secondary analysis included determining the pharmacokinetics of ILB® and measurement of putative biomarkers of ALS progression on weeks 1 (baseline), 5, 10, 24 and 38, including urinary p75 extracellular domain (p75^ECD^) [30] and plasma neurofilament light chain (NfL) [31]. Additional exploratory biomarker analyses were included in the protocol including HGF release following ILB® administration, which is also reported here.

### Sample collection

For ILB® and HGF pharmacokinetic analyses, at least 12 mL of whole blood was collected in anti-coagulant-treated Vacutainer tubes (3.2% sodium citrate) on the first day of ILB® administration 30 minutes prior to injection, as well as 30 minutes and 1, 2, 2.5, 3, 4, and 5 hours post-injection. Vacutainers were inverted 8-10 times and then centrifuged at 2000 x *g* for 10 minutes within 60 minutes of collection at 4°C. Plasma was aliquoted into cryovials and stored at -20°C.

Plasma samples for NfL were collected in one 6 mL EDTA vacutainer, inverted 8-10 times and centrifuged for 10 minutes at 2000 x *g* at room temperature within 60 minutes after collection. Plasma was aliquoted into two cryotubes and stored at -70°C. Where possible the first urine of the morning was also obtained on the same days as the additional blood samples to analyse urinary p75^ECD^ levels. Samples were stored at 4°C until analysis. Samples were taken on weeks 1, 5, 10, 24 and 38 of ILB® treatment (where possible), and two weeks after the end of treatment.

Pre-analytical quality control (QC) indicators collected during sample processing are listed in S4 Tables along with information regarding samples that deviated from the protocolised methods.

All clinical and laboratory staff involved in the collection and preparation of participants’ research samples at the Queen Elizabeth Hospital received Good Clinical Practice (GCP) training and the appropriate laboratory training applicable to GCP and Good Laboratory Practice (GLP). All laboratory equipment used throughout the study were serviced and calibrated (records retained) in accordance with GLP requirements.

### Pharmacokinetic and biomarker analyses

Plasma samples for pharmacokinetic analysis of ILB® were sent to and analysed by Eurofins BioPharma Product Testing, Munich GmbH using a fluorescence probe assay (Heparin Red® Kit; REDPROBES, Germany). Haemolysed samples from four patients were excluded from the analysis, as the autofluorescence of haemoglobin may interfere with the assay [32]. An incomplete pharmacokinetic series led to the exclusion of one further patient. Calibration standards of 10.0, 7.5, 5.0, 2.5, 1.0, 0.5 and 0.0 μg/mL of ILB® were used in triplicates, while internal QC samples of the following concentrations: 6.4, 4.5, 1.8, 0.0 μg/mL were added in sextuplicates (two sets of triplicates in different locations on the plate) to each plate. The imprecision (coefficient of variance) and bias ≤30% was regarded as acceptable (see Supplementary QC data in tables S5A-F). The level of quantification for the assay was set for each plate separately and was defined by restricting the range to calibrators within expected accuracy. If required, patient samples were diluted to bring the ILB® concentration into a range of 7.5 μg/mL – 0.5 μg/mL. Patient samples and internal QC samples were performed in sextuplicates; with the blank, and calibration standards carried out in triplicates.

Plasma samples for pharmacokinetic analysis of HGF were sent to and analysed by Neuregenix Ltd, UK. HGF quantification was carried out using the Quantikine® enzyme-linked immunosorbent assay (ELISA; Cat No: DHG00B, R&D Systems, UK). Patient samples were measured in duplicates at two separate dilutions (1:10 and 1:100) providing four measurements within quantification range for most samples. The assay range was 125 - 8,000 pg/mL with an assay sensitivity of 40 pg/mL. The accepted imprecision (difference between minimum and maximum measurement) and bias for the calibrators was <10% and <±20% respectively, with the total accepted error (TAE) not expected to exceed 30% (assay QC measures are presented in supplementary table S6A-C). No deviations from the analytical protocol were reported. Analyses were conducted in line with the recommendations of the EMA reflection paper for laboratories that perform the analysis or evaluation of clinical trial samples (EMA/INS/GCP/532137/2010) and the OECD Principles of GLP.

Plasma and urine samples for analysis of p75^ECD^ and NfL, respectively, were sent to Neuregenix Ltd, UK. p75^ECD^ was analysed in sample triplicate by Neuregenix laboratories using a commercial quantitative sandwich ELISA purchased from Abcam (Cambridge, UK) and the analysis of NfL was subcontracted to NeuroKemi, Klinisk Kemi, Molndal Hospital, Sweden. The test range for the ELISAs used were 78 pg/mL – 5,000 pg/mL with a sensitivity of 35.6 pg/mL. All laboratory staff involved in the evaluation of research samples received GCP training and the appropriate laboratory training applicable to GCP and GLP to integrate the organisation, procedures, processes, and resources of the laboratory. The laboratory employs a system of internal quality control (IQC) and external quality assurance (EQA) to monitor the quality of test results. This program includes analysis and evaluation of internal quality control samples with every batch (including blanks, duplicate samples, standard reference materials, and enriched samples). All laboratory equipment used throughout the study were serviced and calibrated (records retained) in accordance with GLP requirements.

### Statistical analysis

Due to the nature of the disease and early stage of drug evaluation, no formal sample size calculations were performed. The trial’s size was determined by availably of patients at a single site study, potential risk of exposure to patients, and timing of recruitment to allow results to contribute to the continued drug development.

For the primary outcome, the number of SAEs and AEs, number of intolerable adverse events, total drug administered, number of administrations, number and length of interruptions, and number of discontinuations were calculated per evaluable patient and summarised descriptively. Analyses was performed on the modified intention-to-treat (mITT) principle, in which only patients who received at least one injection of ILB® were analysed. Such patients were defined as being ‘evaluable’. The per-protocol (PP) population is defined as those patients who complete the initial treatment period (the first 10 treatment weeks) without missing a week of therapy. Where appropriate, this population was highlighted during analysis, however no comparison of results from the mITT and PP populations were made.

An initial safety assessment was performed by the data monitoring committee (DMC) after the first two sequential patients were recruited and had received at least four weeks of therapy (sentinel patients). The DMC then met at least three-monthly for the first year and annually thereafter. Pre-specified criteria for stopping the study were:

a. ≥ 1 patient of the first sentinel patients experienced a Serious Adverse Event related to IMP
b. ≥ 33% of patients (with n>3) recruited to the study show a significant decrease in ALSFRS-R score (>50%) compared to baseline during the 10-week dosing period.

Descriptive analyses of the longitudinal ALSFRS-R (symptom) and ALSAQ-40 (QoL) outcomes are presented. The plasma concentrations of ILB® and HGF are presented over time with area under the curve (AUC), maximum concentration (C_max_), time to maximum concentration (T_max_), and half-life (t_1/2_) calculated only for patients whose samples were available for the complete pharmacokinetic series and were of acceptable quality (not haemolysed). Analyses were performed using R version 4.1.0. The pharmacokinetic relationship between ILB® and HGF was calculated using regression analysis (Medcalc).

Finally, a descriptive analysis of urinary p75^ECD^ and plasma NfL concentrations are also presented.

### Trial registration

EudraCT: 2018-000668-28

clinicaltrials.gov: NCT03705390

This trial adheres to the principles of GCP in the design, conduct, recording and reporting of clinical trials as listed in part 2, “Conditions and Principles which apply to all Clinical Trials” under the header “Principles based on Articles 2 to 5 of the EU GCP Directive” in the Medicines for Human Use Clinical Trials Regulations (as amended in SI 2006/1928). For clarity, the study did not conform to all aspects of the International Conference on Harmonisation (ICH) E6 R2 Guidelines for GCP (also known as ‘ICH GCP’). Of note, we did not use an external database, perform 100% source data verification, and only primary outcome data were analysed in parallel by a second, independent statistician.

## Results

Between 18-Apr-2019 and 27-Mar-2020, 11 patients were recruited into the ALS trial of repeated weekly injections of 2 mg/kg ILB® (Fig 1). Patient characteristics are described in Table 1. The median age for patients in the trial was 57 years (range 44 to 77) with the majority being male (7 out of 11). Post-registration, there were no treatment discontinuations or withdrawals.

**Fig 1.**
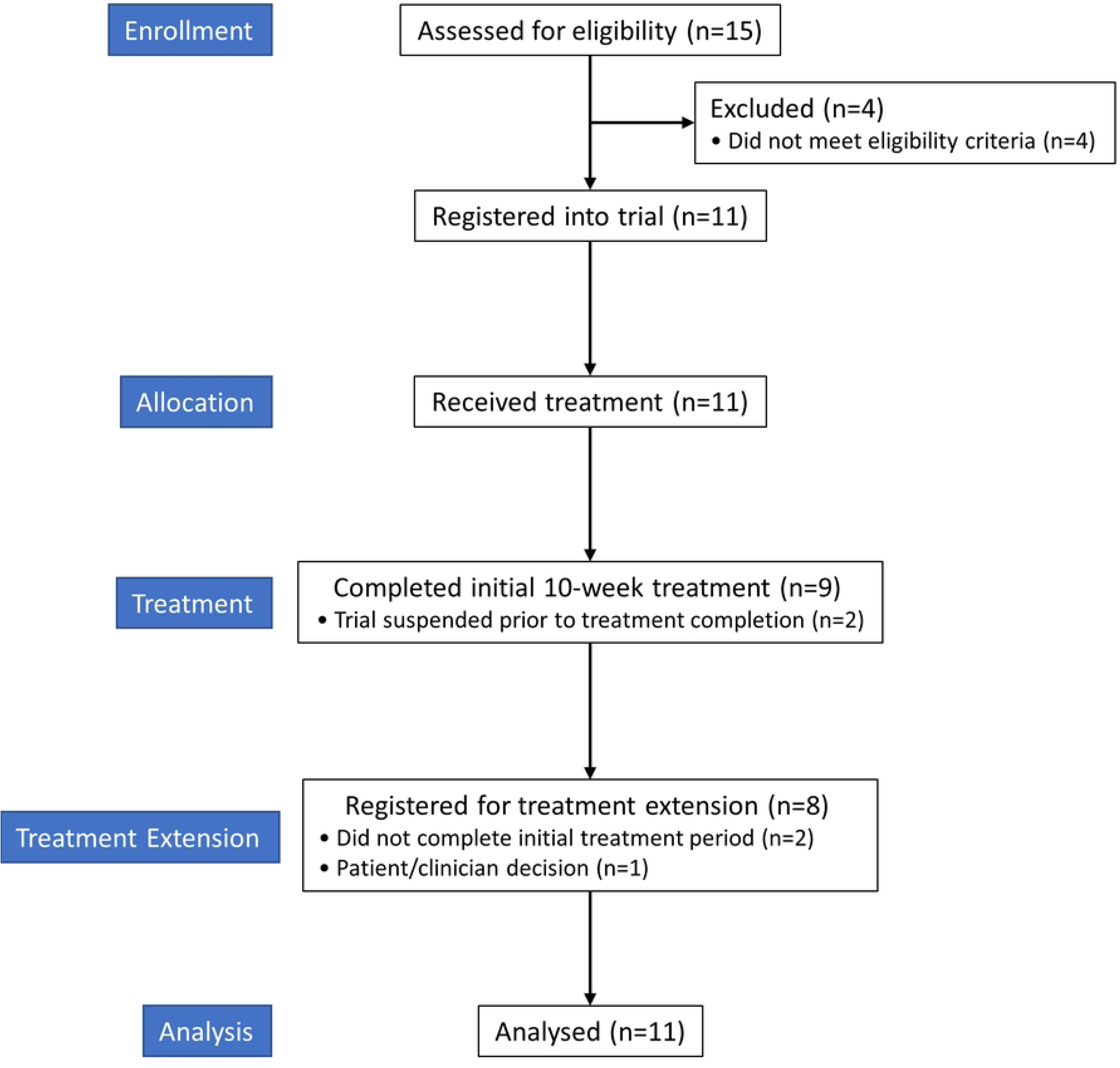
ALS trial profile. CONSORT diagram of the ALS trial.

**Table 1.**
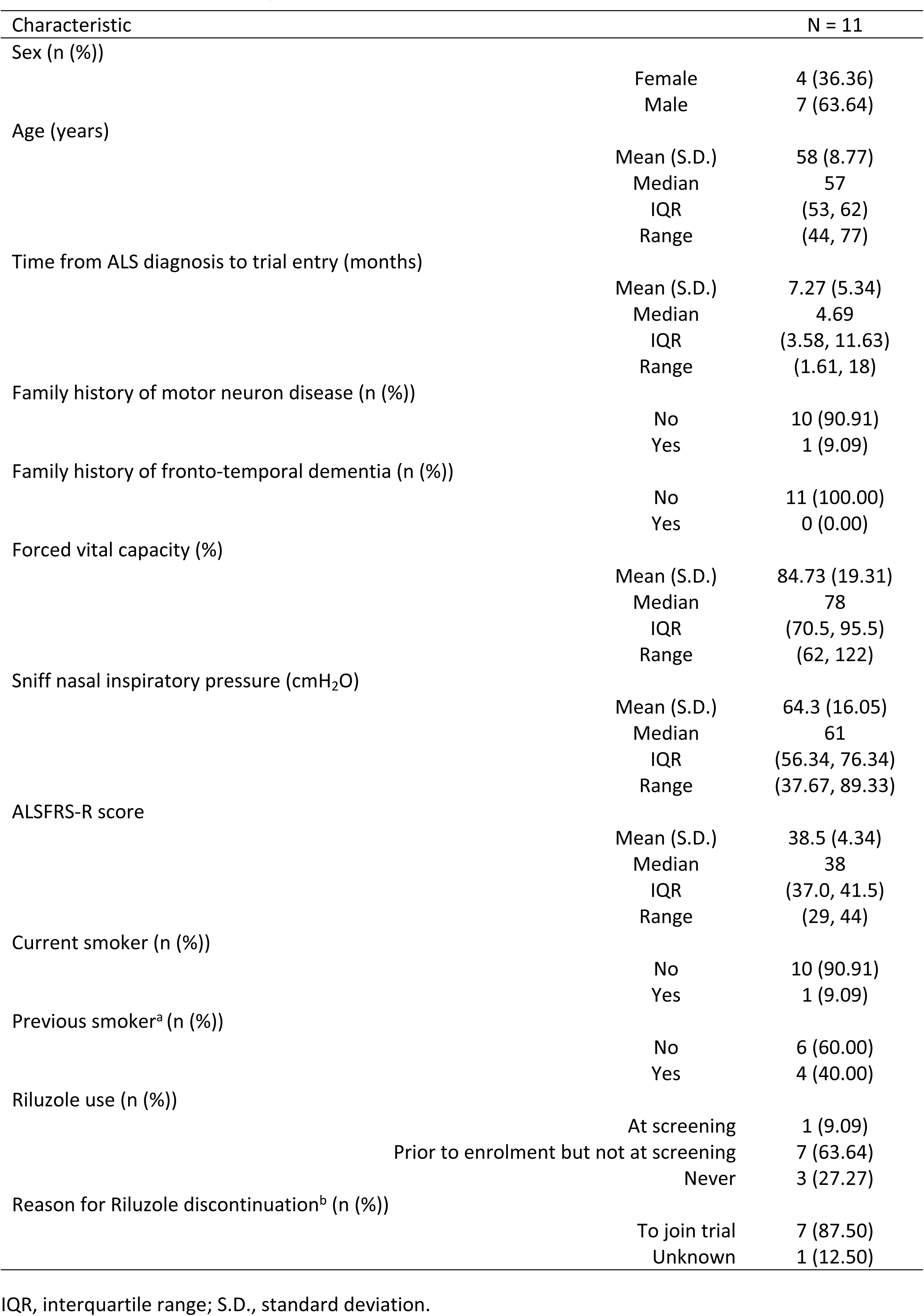
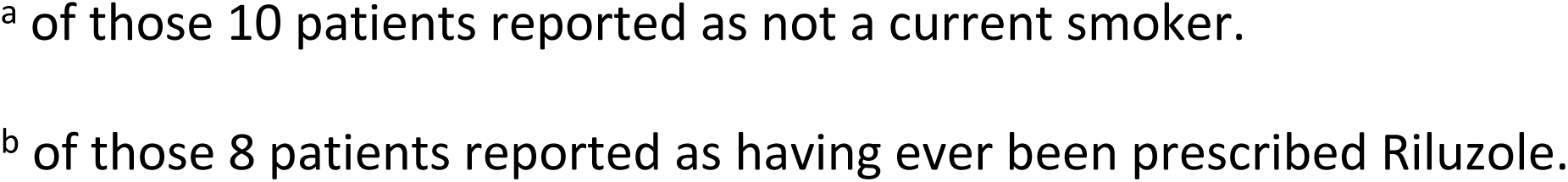
Baseline patient characteristics.

| Characteristic | N = 11 |  |
| --- | --- | --- |
| Sex (n (%)) |  |  |
|  | Female | 4 (36.36) |
|  | Male | 7 (63.64) |
| Age (years) |  |  |
|  | Mean (S.D.) | 58 (8.77) |
|  | Median | 57 |
|  | IQR | (53, 62) |
|  | Range | (44, 77) |
| Time from ALS diagnosis to trial entry (months) |  |  |
|  | Mean (S.D.) | 7.27 (5.34) |
|  | Median | 4.69 |
|  | IQR | (3.58, 11.63) |
|  | Range | (1.61, 18) |
| Family history of motor neuron disease (n (%)) |  |  |
|  | No | 10 (90.91) |
|  | Yes | 1 (9.09) |
| Family history of fronto-temporal dementia (n (%)) |  |  |
|  | No | 11 (100.00) |
|  | Yes | 0 (0.00) |
| Forced vital capacity (%) |  |  |
|  | Mean (S.D.) | 84.73 (19.31) |
|  | Median | 78 |
|  | IQR | (70.5, 95.5) |
|  | Range | (62, 122) |
| Sniff nasal inspiratory pressure (cmH<sub>2</sub>O) |  |  |
|  | Mean (S.D.) | 64.3 (16.05) |
|  | Median | 61 |
|  | IQR | (56.34, 76.34) |
|  | Range | (37.67, 89.33) |
| ALSF<sub>RS</sub>-R score |  |  |
|  | Mean (S.D.) | 38.5 (4.34) |
|  | Median | 38 |
|  | IQR | (37.0, 41.5) |
|  | Range | (29, 44) |
| Current smoker (n (%)) |  |  |
|  | No | 10 (90.91) |
|  | Yes | 1 (9.09) |
| Previous smoker<sup>a</sup> (n (%)) |  |  |
|  | No | 6 (60.00) |
|  | Yes | 4 (40.00) |
| Riluzole use (n (%)) |  |  |
|  | At screening | 1 (9.09) |
|  | Prior to enrolment but not at screening | 7 (63.64) |
|  | Never | 3 (27.27) |
| Reason for Riluzole discontinuation<sup>b</sup> (n (%)) |  |  |
|  | To join trial | 7 (87.50) |
|  | Unknown | 1 (12.50) |

IQR, interquartile range; S.D., standard deviation.
<sup>a</sup> of those 10 patients reported as not a current smoker.
<sup>b</sup> of those 8 patients reported as having ever been prescribed Riluzole.

As a result of the COVID-19 pandemic and the high-risk status of study participants, recruitment and study treatment was suspended with a decision made by the funders and sponsor to terminate the study prematurely in October 2020. A single point long-term remote follow-up visit was undertaken, for which each patient was asked to provide consent for a visit within 2021. Additional QoL data were collected along with information on all medications each patient was taking. Eight patients attended their long-term follow-up visits between 21-Jun-2021 and 28-Jul-2021. The remaining three patients had died since their previous visit: one at 56 weeks, one at 62 weeks; and a third at 53 weeks after receiving their last ILB® dose. Causes of death were unknown. Median follow-up (defined as length of time from randomisation to date last seen) for all patients was 21.2 months (range 14.3 to 25.7).

After four weeks of treatment and following a review by the DMC of the initial two sentinel patients recruited, no patient or trial specific safety stopping rules were met. Following amendments to the trial protocol and subsequent reviews by the UK’s competent authority (MHRA) and the trial’s research ethics committee, recruitment was extended, and further treatments were permitted. A total of 11 patients were recruited to receive the initial 10 weeks of treatment, they then had the option to extend treatment to 24 weeks, with the opportunity to further extend to up to 48 weeks of treatment (in total, including the initial and first extension treatment periods). Eight patients enrolled into the treatment extension, one declined, and two had not completed the initial 10 weeks of treatment prior to the trial being stopped. Six out of the eight patients enrolled had a minimum of 24 weeks treatment. This treatment schedule is summarised in Fig 1 with treatment delivery details for each patient shown in Fig 2. The quantity of study drug administered is summarised in Table 2, with detailed ILB® doses available in S7 Tables.

**Fig 2.**
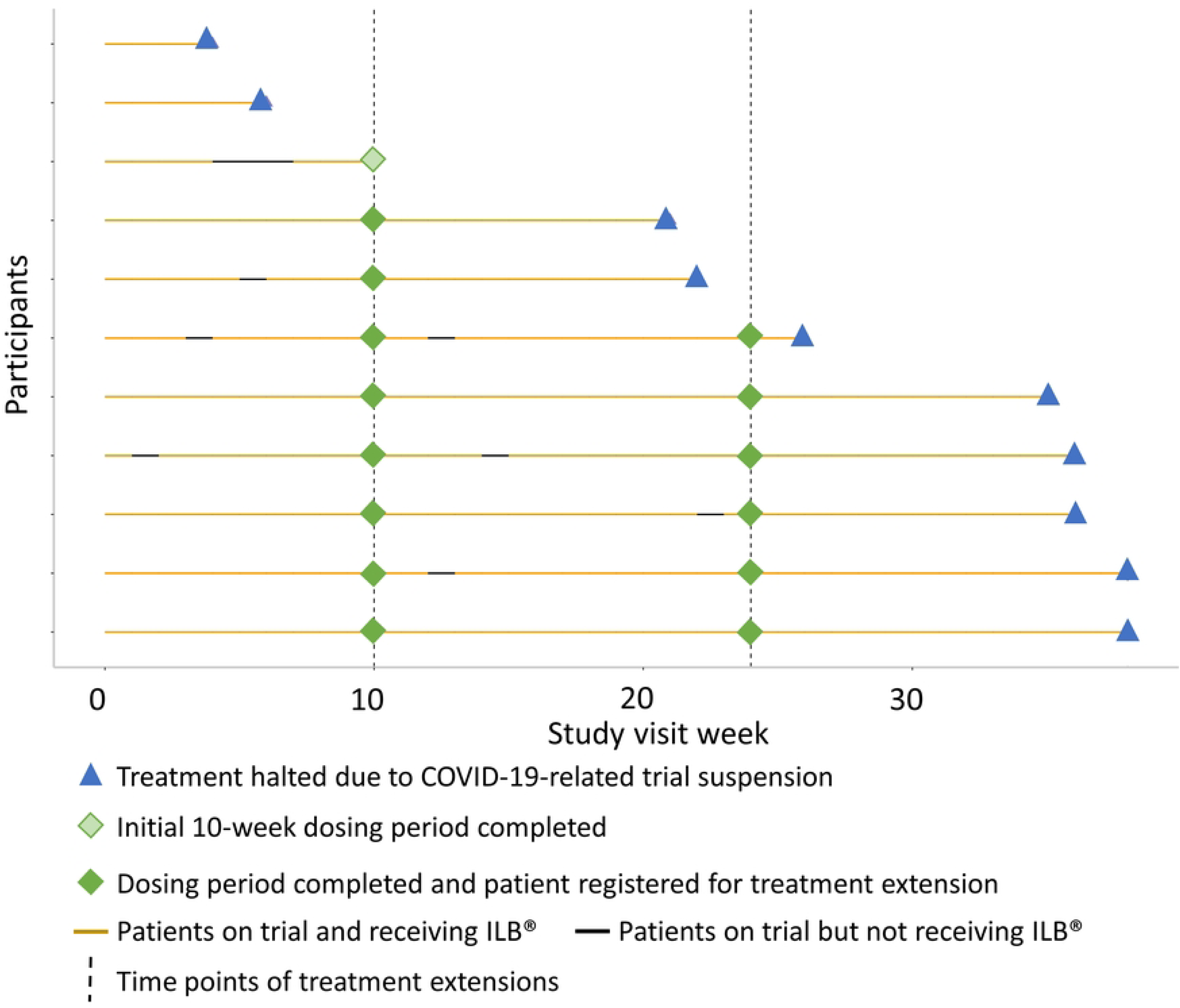
Patients’ treatment during the ALS trial. Swimmer plot to visualise adherence to trial treatment and ordered by patients’ treatment duration.

**Table 2.** Quantity of study drug administered ordered by patients’ treatment duration.

| Number of Treatment Weeks | Number of Treatment Administrations | Average Weight (kg) | Average Dose (mg) | Cumulative Dose (mg) | Number of Interruptions | Duration of Interruption* (weeks) |
| --- | --- | --- | --- | --- | --- | --- |
| 38 | 38 | 86.20 | 172.39 | 6551 | 0 | - |
| 38 | 37 | 90.11 | 180.22 | 6668 | 1 | 1 |
| 36 | 35 | 88.90 | 177.77 | 6222 | 1 | 1 |
| 36 | 34 | 72.08 | 143.62 | 4883 | 2 | 2 |
| 35 | 35 | 75.62 | 151.06 | 5287 | 0 | - |
| 24 | 24 | 87.45 | 175.00 | 4200 | 2 | 2 |
| 22 | 21 | 70.63 | 141.29 | 2967 | 1 | 1 |
| 21 | 21 | 66.43 | 132.81 | 2789 | 0 | 0 |
| 10 | 7 | 59.57 | 119.14 | 834 | 3 | 3 |
| 6 | 6 | 84.80 | 169.33 | 1016 | 0 | - |
| 4 | 4 | 93.20 | 186.25 | 745 | 0 | - |

\* Each interruption lasted 1 week and may or may not have been consecutively. Timing of interruptions can be visualised within Fig 2 where missed treatment weeks are shown as red "X"s.
Note: Each line presents an individual patient.

One serious adverse event (SAE) was reported in one patient during the trial; grade 3 (severe) muscle weakness, which was not related to ILB® and resolved without sequalae. During the trial no DILI were reported, however, one patient did experience raised liver function tests. Details of these measurements are shown in S8 Table. This patient had abnormal LFTs prior to treatment, which persisted and became worse during the 12 weeks of ILB® treatment. In addition, ILB® was not given to this patient on weeks 5, 6 and 7, this was due to no attendance at clinic (visit 5) and the patient experiencing pneumonia, which occurred during the expected visit 6 and 7. Upon investigation of this patient’s laboratory values they were found not to fulfil the relevant criteria for DILI i.e., ALT (>3× ULN persisting for 4 weeks) as stated in S3 Table. Administration of ILB® was not withheld from the patient due to intolerability at any time.

Two hundred and seventy AEs were reported from the 11 participants during the ALS trial (S9 Table). The most common side effect was grade 1 (mild) bruising, which was experienced by nine patients. The majority of events were grade 1/mild (265 (98.1%)), with four (1.5%) grade 2/moderate events, and one (0.4%) grade 3/severe event (Table 3). Of these, 92 (33.4%) AEs were assigned as definitely related to ILB® (bruising), one (0.4%) probably related (discomfort at injection site when touched), and four (1.5%) possibly related (elevated partial thromboplastin time, increased aspartate aminotransferase, increased alkaline phosphatase, and increased alanine aminotransferase). All were grade ≤2 (mild/moderate). Therefore, no intolerable AEs were reported during the ALS trial.

**Table 3.** Number of adverse events experienced by patients ordered by patients’ treatment duration.

| Number of Treatment Weeks | Number of Treatment Administrations | Number of Events as Defined by CTCAE v4.0* |  |  | Total Number of Events |
| --- | --- | --- | --- | --- | --- |
|  |  | Grade 1 | Grade 2 | Grade 3 |  |
| 38 | 38 | 44 | 1 | 1 | 46 |
| 38 | 37 | 19 | 0 | 0 | 19 |
| 36 | 35 | 23 | 0 | 0 | 23 |
| 36 | 34 | 33 | 2 | 0 | 35 |
| 35 | 35 | 48 | 0 | 0 | 48 |
| 26 | 24 | 14 | 0 | 0 | 14 |
| 22 | 21 | 25 | 0 | 0 | 25 |
| 21 | 21 | 29 | 1 | 0 | 30 |
| 10 | 7 | 14 | 0 | 0 | 14 |
| 6 | 6 | 6 | 0 | 0 | 6 |
| 4 | 4 | 10 | 0 | 0 | 10 |

\* According to National Cancer Institute (NCI) Common Terminology Criteria for Adverse Events (CTCAE) v4.03 (2010). Available from:
https://ctep.cancer.gov/protocoldevelopment/electronic_applications/ctc.htm#ctc_40 where grades 1-to-3 are defined as:
- Grade 1 – mild; asymptomatic or mild symptoms; clinical or diagnostic observations only; intervention not indicated. - Grade 2 – moderate; minimal, local, or non-invasive intervention indicated; limiting age-appropriate instrumental activities of daily life. - Grade 3 – severe or medically significant but not immediately life-threatening; hospitalisation or prolongation of hospitalisation indicated; disabling; limiting self-care activities of daily life.
Note: Each line presents an individual patient.

No patient experienced a 50% deterioration in ALSFRS-R (symptom) or ALSAQ-40 (QoL) scores that were predefined individual stopping criteria of the trial. Furthermore, in all patients, ALSFRS-R and ALSAQ-40 scores appeared stable, with no clinically significant changes (Fig 3). One patient exhibited a greater deterioration when their ALSFRS-R score prior to first treatment was compared to that at the end of treatment (Table 4). However, their end of treatment ALSFRS-R score was not appreciably different from their score at trial registration.

**Fig 3.**
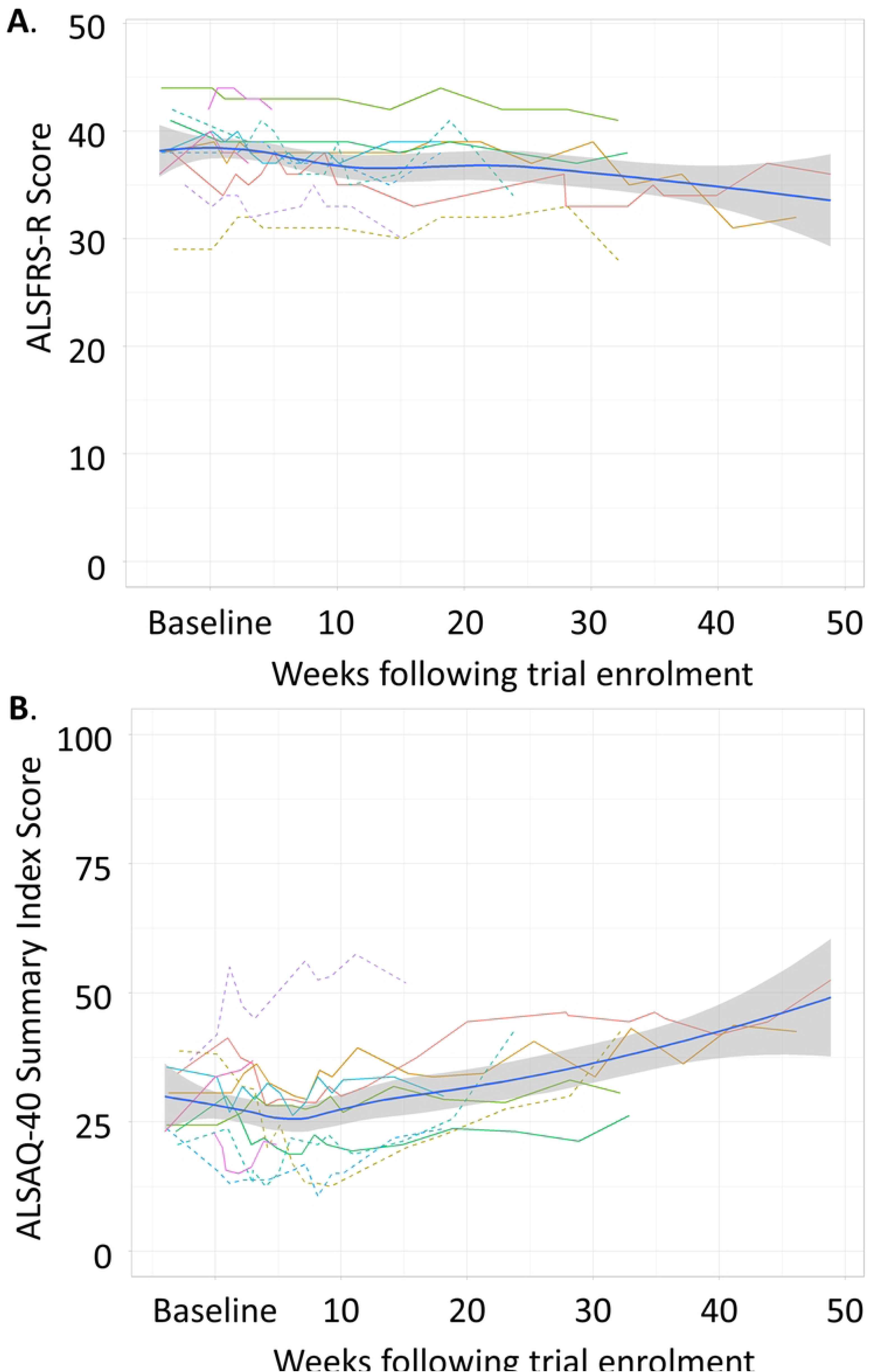
Symptom progression and quality of life of patients during the ALS trial. Trajectories of ALSFRS-R scores (A) and ALSAQ-40 Summary Index scores (B) through time during the ALS trial, with trend lines (royal blue) and ± 1 standard error (grey). Patients’ data are presented as the modified intension-to-treat population, with the solid line showing those who are also in the per-protocol population. Each patient is presented as a different coloured line.

**Table 4.** Patients’ disease progression prior to and during the ALS trial ordered by patients’ treatment duration.

| Number of Treatment Weeks | Number of Treatment Administrations | ALSFRS-R score at trial registration | ALSFRS-R score at the beginning of treatment | Functional change pre-treatment<sup>&</sup> | ALSFRS-R score after 10 weeks of treatment | Functional change during first 10 weeks of treatment<sup>&</sup> | ALSFRS-R score at end of treatment | Functional change during trial<sup>&</sup> |
| --- | --- | --- | --- | --- | --- | --- | --- | --- |
| 38 | 38 | 38 | 39 | -1.07 | 38 | 0.48 | 32 | 0.67 |
| 38 | 37 | 38 | 34 | 4.43 | 35 | -0.48 | 36 | -0.18 |
| 36 | 35 | 44 | 44 | 0.00 | 43 | 0.49 | 41 | 0.41 |
| 36 | 34 | 29 | 29 | 0.00 | 31 | -0.98 | 28 | 0.14 |
| 35 | 35 | 41 | 39 | 2.21 | 39 | 0.00 | 38 | 0.14 |
| 26 | 24 | 42 | 40 | 2.14 | 39 | 0.48 | 34 | 1.14 |
| 22 | 21 | 38 | 38 | 0.00 | 37 | 0.48 | 38 | 0.00 |
| 21 | 21 | 38 | 40 | -2.14 | 38 | 0.95 | 39 | 0.24 |
| 10 | 7 | 35 | 33 | 4.00 | 33 | 0.00 | 30 | 0.87 |
| 6 | 6 | 44 | 42 | 4.14 | - | - | 42 | 0.00 |
| 4 | 4 | 36 | 40 | -4.29 | - | - | 37 | 4.14 |

<sup>&</sup> Data presented as change in ALSFRS-R (symptom) score defined as point change per month
Note: Each line presents an individual patient.

Pharmacokinetics of ILB® confirmed detectable plasma ILB® with a median half-life of 7.2 hours (IQR: 5.1, 10.9) (Fig 4 and S10 Tables). Pharmacokinetics of plasma HGF also confirmed detectable systemic HGF release following ILB® administration with a strong significant relationship between ILB and HGF concentration in the samples (Fig 5 and S11 Tables). C_max_ levels were similar to those seen in the previously reported ILB® study [25] but high levels of circulating HGF persisted for longer (t_1/2_ is beyond the six-hour time point). The positive correlation between the concentration of ILB® and HGF in patients’ plasma observed (R^2^=84.64%) confirmed previous reports of an ILB® dose-dependent increase in plasma HGF [22–25] (Fig 5B).

**Fig 4.**
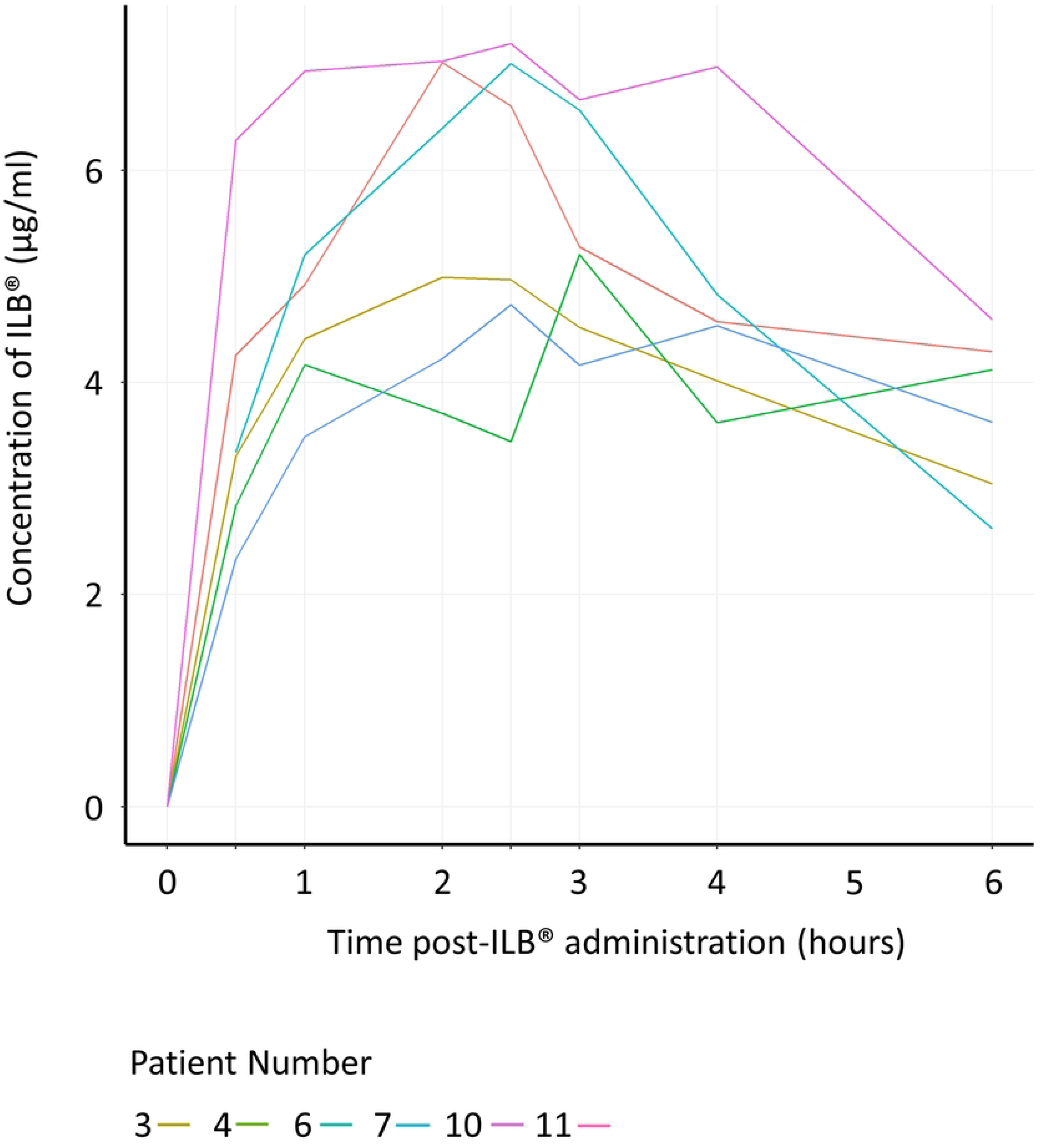
Pharmacokinetics of ILB®. Repeated measures of the plasma concentration of ILB® per patient are shown. Plasma samples were analysed before, and 0.5, 1.0, 2.0, 2.5, 3.0, 4.0, 6.0 hours after initial ILB® injection via a fluorescence probe assay (Heparin Red® Kit). Note: Each patient with a complete pharmacokinetic sample series is presented as a different coloured line. Results from four patients were not included due to haemolysis of the plasma in some of their samples. In addition, the results from one patient’s samples were not included due to an incomplete pharmacokinetic series. The sample prior to ILB® administration for one patient was not analysed/missing but all other samples were collected and have, therefore, been included.

**Fig 5.**
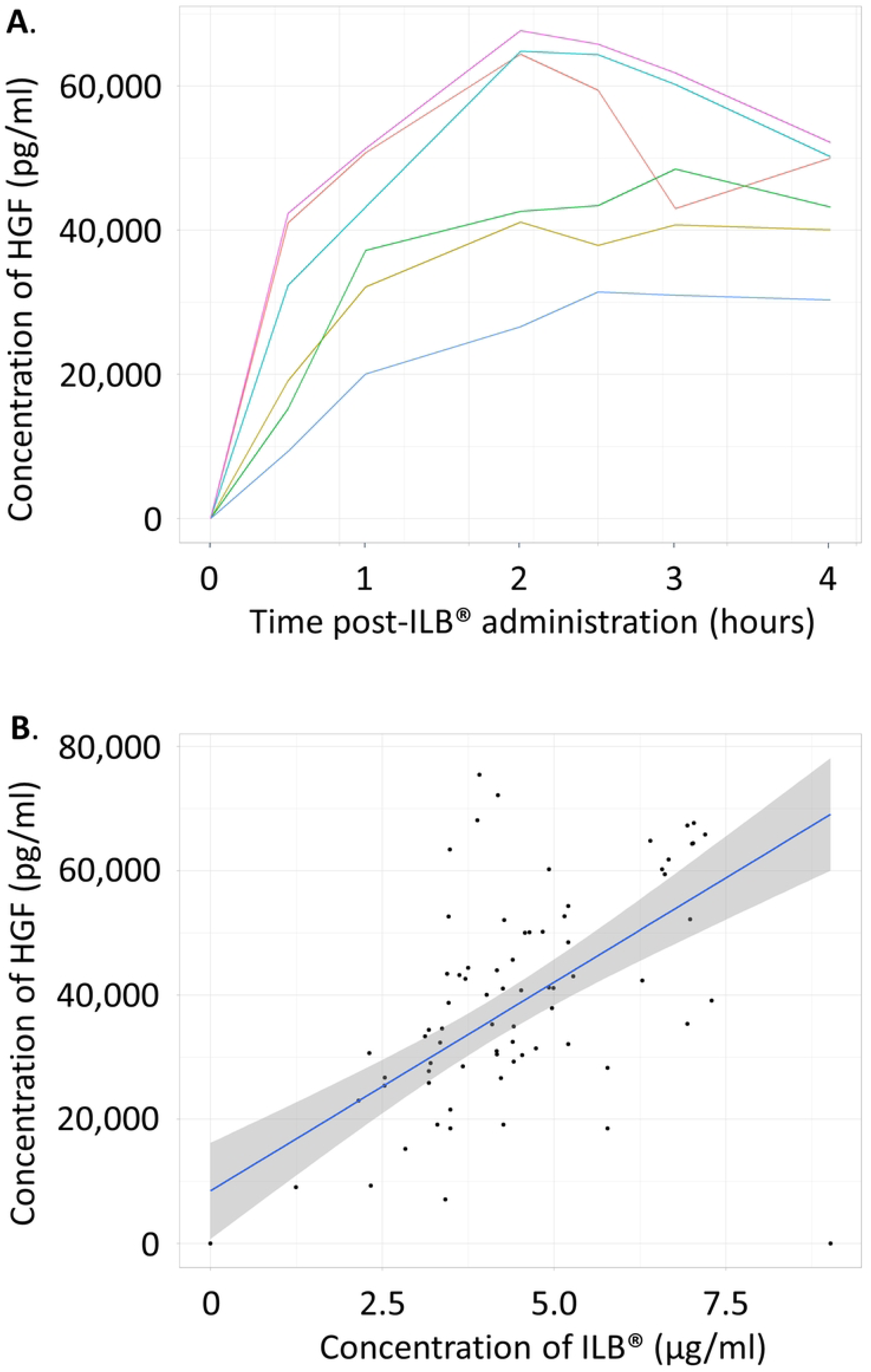
Pharmacokinetics of HGF and relationship with ILB®. Repeated measures of the plasma concentration of human growth factor (HGF) per patient are shown (A). Plasma samples were analysed before, and 0.5, 1.0, 2.0, 2.5, 3.0, 4.0, 6.0 hours after initial ILB® injection *via* Quantikine® ELISA immunoassay. Concentrations of plasma HGF and ILB® measured at the same time points were plotted against each other to assess their relationship (B). Note: Each patient with a compete pharmacokinetic sample series is presented as a different coloured line. The results from four patients were not included due to haemolysis of the plasma in some of their samples. In addition, one patient’s sample data were not included due to an incomplete pharmacokinetic series. The sample prior to ILB® administration for one patient was not analysed/missing but all other samples were collected and have, therefore, been included.

Duration of treatment and follow-up differed for each patient; therefore, for those sample time points collected, the biomarkers of ALS progression (urinary p75^ECD^ and plasma NfL levels) were analysed and are shown in Fig 6. In general, these data suggest that there was no change in these measured disease biomarkers in treated patients during the 38-week trial.

**Fig 6.**
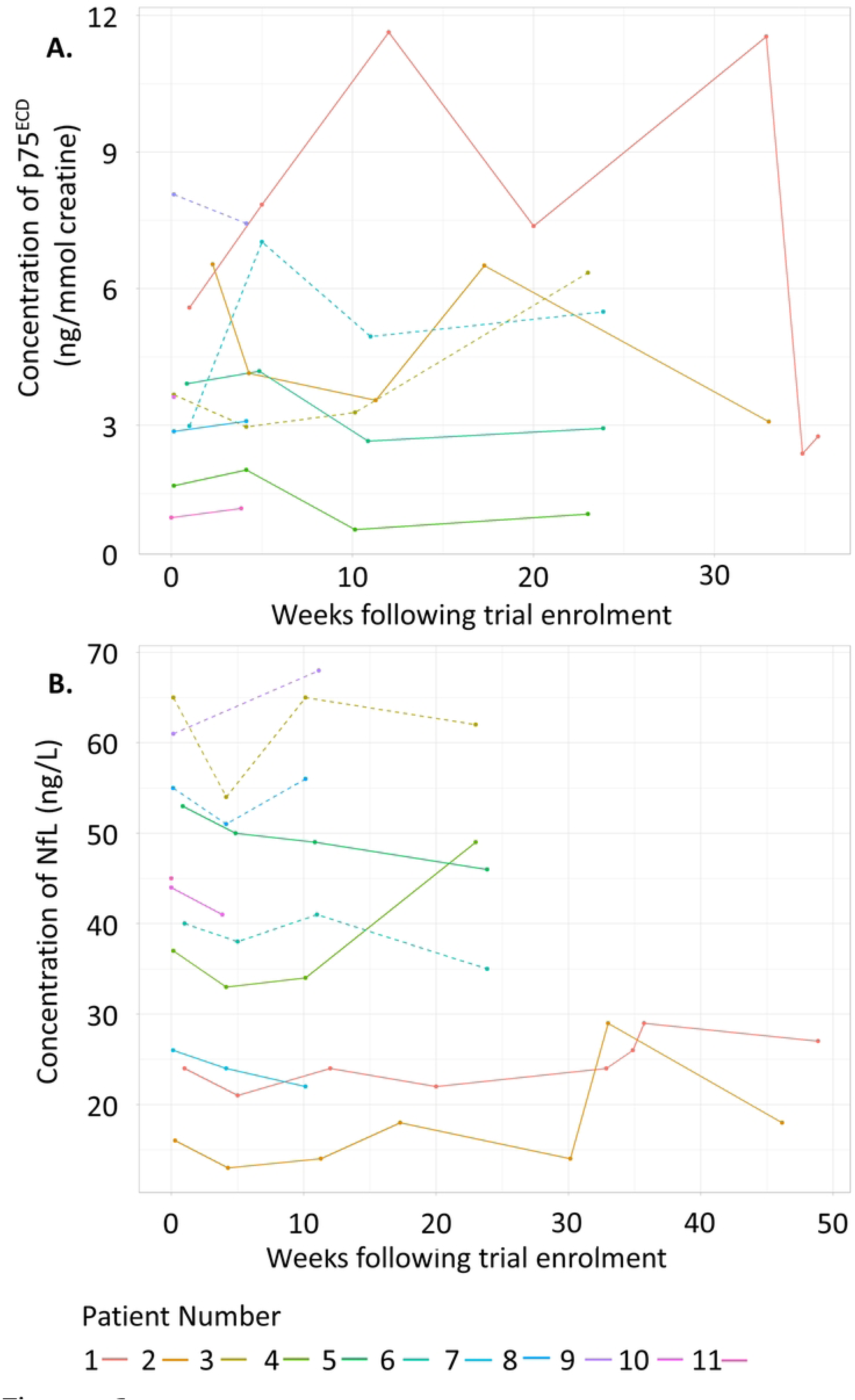
Biomarkers of ALS progression. Where available, concentrations of the disease progression markers, urinary p75 extracellular domain (p75^ECD^) (A) and plasma neurofilament light chain (NfL) (B), were measured *via* ELISA. Patients’ data are presented as the modified intension-to-treat population, with the solid lines those who are also in the per-protocol population. Each patient is presented as a different coloured line.

## Discussion

The ALS trial was a prospective, single-arm, open-label phase II clinical trial in patients with a definitive diagnosis of ALS based on El-Escorial criteria to determine the safety and efficacy of long-term weekly dosing of a higher concentration of a sulphated form of LMW-DS (ILB®) than had been previously tested [25]. Recruitment of 15 patients was planned for treatment up to 10 weeks in the first instance extending to up to 24 and 48 weeks following a favourable toxicity profile as assessed by the treating clinician, provided no patient specific stopping rules had been met, and provided patients were willing to proceed. However, due to the COVID-19 pandemic, the high-risk status of trial patients, and the frequent hospital-based visits, both recruitment to the trial and further treatment was suspended after 11 patients had been recruited. As a result, patients were treated with interrupted and varying numbers of weekly ILB® treatments, ranging from 4 to 38. There were no treatment discontinuations within the initial 10-week dosing schedule, no withdrawals of consent, and no patient experienced a dose interruption of greater than three weeks.

During the trial 270 AEs were reported in all 11 patients, but overwhelmingly these were mild (265 grade 1 AEs), with no AEs deemed at least possibly related to treatment graded moderate or higher. Reassuringly, this was analogous to results from the pilot clinical study of shorter term administration of 1 mg/kg ILB® in ALS patients, where the most frequent AEs were related to subcutaneous injection site bruising with no major bleeding or haematoma formations [25]. There was one SAE reported during and one episode of elevated liver function tests during this ALS trial, but upon investigation neither were attributed to the trial IMP. While there were three reported deaths in the patient cohort, all of these happened after early termination of the trial due to the COVID-19 pandemic. All deaths occurred at least 53 weeks after the patients received their final ILB® dose and none were attributed to ILB®. Given this favourable safety profile, and the quantity of study drug administered, we conclude that ILB® was safe and tolerable at this higher dose and frequency, and therefore, the primary outcomes of the trial were met.

ALSFRS-R and ALSAQ-40 were used to assess the patient function and disease progression before, during, and after ILB® administration. The ALSFRS-R has been widely used in MND trials to ascertain the rate of progression of the disease and is a predictor of disease progression. A single point reduction of ALSFRS-R has been shown to increase risk of death, or need for respiratory support by 7% [33]. It has also been shown that slowing the decline of the score correlates with prolonging median survival [34], with a 20% reduction of the progression rate regarded as clinically relevant [35]. The ALSFRS-R score changed minimally during the 38-week trial of ILB® treatment. The mean rate of ALS progression within the ProACT database is reported as a 1.02 decrease per month [36]. While not designed to show an effect on ALSFRS-R, this trial may indicate a slowing of disease progression in this ILB® treated patient cohort, although the limitation of its open-label design is noted.

The ALSFRS-R data reported in the previous ILB® trial of five weekly doses of 1 mg/kg suggested functional recovery within one week of treatment initiation, which increased to statistical significance during the treatment period [25]. This current trial showed no such clear-cut functional benefit to patients. However, there were significant differences between these two trials. The most notable difference was the ILB® dose used and length of treatment. While the previous trial used 1 mg/kg for five continuous weeks, the current trial used 2 mg/kg dose for a highly variable length of time (due to the COVID restrictions that came into effect during the trial). Moreover, the inclusion criteria of this current trial were more stringent (definite diagnosis of ALS according to El Escorial Criteria required) than the previously reported clinical trial [25] leading to a less heterogenous patient population in this current trial. Therefore, while long-term weekly dosing of 2 mg/kg ILB® was not powered to detect efficacy-related measures, the ALSFRS-R measures of patients indicated a slowing of disease progression during treatment (relative to pre-enrolment disease progression) in 7 of the 11 patients.

The QoL score, ALSAQ-40, is a reliable and validated method for measuring QoL in patients with MND and could be used to measure the treatment outcome of drug therapies for ALS/MND [29, 37, 38]. Similar to ALSFRS-R, even a small change in the score can reflect adverse effects on patient QoL [39]. During this trial, like ALSFRS-R, the ALSAQ-40 score changed minimally, which may also be an indication of slowing of disease progression, although definitive conclusions regarding these data cannot be drawn as the trial was not powered to detect this.

The optimal clinical dose of ILB® remains to be determined. Theoretical modelling together with in vitro and animal data have demonstrated a non-linear dose response with ILB® [21], an observation that should be considered in future clinical trial design and when evaluating the relative outcome of this clinical trial *versus* the previously reported ALS trial with ILB® [25].

Urinary p75^ECD^ gets cleaved from cell membranes following cell injury with levels found to be higher in patients with ALS, which increase as the disease progresses [30]. More recently, a meta-analysis also found p75^ECD^ could be a potential biomarker and indicator of progression of ALS [40]. Furthermore, as a marker of axonal damage/degeneration, potential use of plasma NfL as a diagnostic and prognostic marker of ALS within clinical trials has been suggested [41, 42]. During this ALS trial, sample collection to detect p75^ECD^ and NfL was variable primarily due to differences in the duration of treatment and follow-up for each patient. Consequently, although detected levels of both metabolites did not change significantly during the trial, due to the considerable data heterogeneity, no conclusions could be drawn about the significance of the biomarker data.

The pharmacokinetic data indicate that, as seen previously, HGF is released from the endothelial bed by ILB® [21]. The maximal HGF release seen with the 2 mg/kg dose is not higher than that seen with 1 mg/kg ILB®, which suggests that 1 mg/kg ILB® is able to release almost all available HGF from the extracellular matrix of the vascular bed and suggesting that increasing the ILB® dose has no benefit in terms of increasing the release of growth factors. It is, however, important to note that the higher concentrations of ILB® may have sequestered the released HGF in the plasma for longer, thus limiting its bioavailability.

The pharmacokinetic evaluation was hampered due to poor quality blood samples, and this draws attention to the need of stringent QC at both the preanalytical and analytical stages of clinical trial samples. This problem was mitigated here by rejecting compromised samples that were easily identified. It is recognised that poor quality samples should be replaced at the pre-analytical stage, while at the analytical stage attention should be given to the total error in the measurements and the correct analysis of internal QC samples. It is recommended that the quality performance of laboratories collecting, storing, and analysing samples be fully documented before clinical trial samples are analysed.

The primary objective of this trial was safety with the secondary outcomes of patient function and disease progression based on patient reported outcomes and biomarkers. Lung function was measured by FVC during screening, at various visits during treatment (weeks 6, 14, 19, 24, 28, 33, and 38), and at the end of treatment. However, these measurements were used only as an indicator of disease progression for the clinician to check eligibility as the trial was small in terms of patient numbers and was not designed or powered to assess efficacy.

In summary, this ALS trial showed that long-term weekly injection of 2 mg/kg ILB® was safe and well tolerated in a cohort of patients with confirmed ALS. A larger trial with longer follow-up, optimised dosing and a formal comparator would be needed to make definitive conclusions about efficacy.

## Contributors

VS: Conceptualization; Methodology; Funding Acquisition; Project Administration; Writing – Original Draft Preparation; Writing – Review & Editing; VH: Methodology; Project Administration; Formal Analysis; Data Curation; Writing – Original Draft Preparation; Writing – Review & Editing; DB: Funding Acquisition; Project Administration; Writing – Original Draft Preparation; Writing – Review & Editing; ACJ: Project Administration; Data Curation; Writing – Review & Editing; SJ: Project Administration; Data Curation; Writing – Review & Editing; CP: Project Administration; Data Curation; Writing – Review & Editing; KB: Conceptualization; Funding Acquisition; Writing – Review & Editing; AL: Conceptualization; Funding Acquisition; Writing – Review & Editing; DS: Conceptualization; Methodology; Project Administration; Writing – Review & Editing; LB: Writing – Review & Editing; ZN: Methodology; Writing – Review & Editing; SPB: Conceptualization; Methodology; Funding Acquisition; Writing – Original Draft Preparation; Writing – Review & Editing.

## Data Availability

Participant data and the associated supporting documentation will be available within six months after the publication of this manuscript. Details of our data request process is available on the CRCTU website. Only scientifically sound proposals from appropriately qualified research groups will be considered for data sharing. The decision to release data will be made by the CRCTU Director’s Committee, who will consider the scientific validity of the request, the qualifications and resources of the research group, the views of the Chief Investigator and the trial steering committee, consent arrangements, the practicality of anonymising the requested data and contractual obligations. A data sharing agreement will cover the terms and conditions of the release of trial data and will include publication requirements, authorship and acknowledgements and obligations for the responsible use of data. An anonymised encrypted dataset will be transferred directly using a secure method and in accordance with the University of Birmingham’s IT guidance on encryption of data sets.

## Acknowledgements

We thank the patients who took part in the trial; the Queen Elizabeth University Hospital Birmingham Neurology research nursing team (Birmingham, UK); the staff from the CRCTU, University of Birmingham including Dr Siân Lax for their contributions to the paper. We would also like to acknowledge the contribution of the independent Data Monitoring Committee (Dr Simon Ellis - Consultant Neurologist; Dr Anthony Thomas - Consultant Neurologist, independent clinician; Elena Frangou - Independent Statistician). We would also like to acknowledge Eurofins BioPharma Product Testing, Munich GmbH who performed the ILB® measurements and Neuregenix Ltd who performed the biomarker analyses.

